# Decline, and Regional Disparities, in Medical Cocaine Usage in the United States

**DOI:** 10.1101/2020.08.25.20181065

**Authors:** Youngeun C. Armbuster, Brian N. Banas, Kristen D. Feickert, Stephanie E. England, Erik J. Moyer, Emily L. Christie, Sana Chughtai, Tanya J. Giuliani, Rolf U. Halden, Kenneth L. McCall, Brian J. Piper

**Author notes:** designated authors contributed equally (YCA, BNB, KDF).

## Abstract

**Purpose:** Cocaine is a stimulant with a complex history that is used in otorhinolaryngological surgeries as a local anesthetic and vasodilator. There is extensive regulation in the United States for the storage and disposal of this Schedule II drug, potentially incentivizing health care professionals to avoid use. This descriptive study characterized medical cocaine use in the United States.

**Methods:** Retail drug distribution from 2002-2017 in units of grams of weight was extracted for each state from the Drug Enforcement Administration’s Automation of Reports and Consolidated Orders System database, which reports on medical, research, and analytical-chemistry use. The percent of buyers (hospitals, pharmacies, providers) was obtained. Use per state, corrected for population, was determined. Available data on cocaine use, as reported by the Medicare and Medicaid programs for 2013 – 2017, also were examined.

**Results:** Medical cocaine use in the US, measured on the basis of mass, decreased 62.5% from 2002 to 2017. Hospitals accounted for 84.9% and practitioners for 9.9% of cocaine distribution in 2017. The number of pharmacies nationwide carrying cocaine dropped by 69.4% to 206. The percent of all US hospitals, practitioners, and pharmacies that carried cocaine in 2017 was 38.4%, 2.3%, and 0.3%, respectively. There was a seven-fold difference in distribution per state in 2002 (South Dakota = 76.1 mg/100 persons, Delaware = 10.1 mg/100 persons). Similarly, there was a ten-fold regional disparity observed for 2017. Relative to the average state, those reporting the highest values (Montana = 20.1 and North Dakota = 24.1 mg/100 persons), were significantly elevated. Cocaine use within the Medicare and Medicaid programs was negligible.

**Conclusion:** Medical cocaine use across the United States exhibited a pronounced decline over a fifteen-year period. If this pattern continues, licit cocaine will soon become an obscure pharmacological relic of interest only to analytical chemists and medical historians.

Key Points
1. **Question:** How has medical use of cocaine, a local anesthetic and vasoconstrictor administered for otorhinolaryngological surgeries and some diagnostic procedures, changed in the United States?
2. **Findings:** Cocaine usage, as reported to the Drug Enforcement Administration has undergone a pronounced (62.5%) decline over the past fifteen-years, while some pronounced regional differences were noted.
3. **Meaning:** Although cocaine has played a key role in the history of anesthesia, the development of safer and non-controlled alternatives may continue to supplant this agent in contemporary use.

---

Koller’s landmark report on cocaine as a local anesthetic was rated as the second most important discovery in the history of anesthesia.^1,2^ Although cocaine was often a preferred topical anesthetic, due to increasing costs, extensive regulations, and habit-forming potential,^3,4^ replacement compounds have been sought.^5^ The US Drug Enforcement Administration (DEA) currently classifies cocaine as a Schedule II drug, which indicates a high potential for abuse but allows for administration for medical purposes, albeit with severe restrictions.^6^ Cocaine can be employed for a variety of purposes in contemporary medicine, including as an anesthetic in various types of surgery as well as diagnosing specific diseases such as Horner syndrome.^7, 8^ The administration of small doses of intranasal liquid cocaine (4% cocaine) is used by otorhinolaryngological surgeons for local anesthesia. Intranasal anesthetics, including cocaine, are absorbed systemically. Use of cocaine as a local anesthetic also results in vasoconstriction of the coronary arteries via stimulation of alpha-adrenergic receptors.^9^ In pregnant females, cocaine can be distributed readily across the placenta.^10^ Recreational usage of cocaine in extreme doses has been associated with myocardial infarction, but this association is not observed with controlled doses used in topical anesthetics.^9^ Chronic recreational usage of cocaine can cause lesions of the nose, sinus, and palate as well as nasal septum perforations.^11^ These potential adverse effects may disincentivize health care providers from medical use of cocaine.^11^

The objective of this report was to quantify the trends in licit cocaine distribution in the United States using DEA data from 2002-2017, and to determine the usage of medical cocaine in the Medicaid and Medicare part D programs from 2013-2017. Based on research with other controlled substances,^12-17^ we hypothesized that cocaine use may be declining and vary regionally across the US.

## METHODS

### Procedures

Retail distribution of cocaine (9041L) by grams weight, number of buyers reported as hospitals, pharmacies, teaching institutions, providers (i.e., surgical centers), and mid-level providers from 2002-2017 in each state was extracted from Automation of Reports and Consolidated Orders System (ARCOS) database (https://www.deadiversion.usdoj.gov/arcos/). The 9041L encompasses various formulations of cocaine hydrochloride including as a monotherapy (4% or 10%) or with adrenaline (Supplemental Table 1).^18^ The year 2017 was the most recent available data set when this analysis was completed (June, 2020). This comprehensive data source captures medical and research use and cocaine distribution as an internal standard for analytical chemistry procedures. ARCOS is based on manufacturers and distributors reporting their controlled substances transactions to the DEA and has been employed in prior pharmacoepidemiology reports of opioids^12-16^ and stimulants.^17^

Medical use of cocaine was also obtained from the Medicare and Medicaid programs from 2013-2017. Medicare Part D Prescriber Data from Centers for Medicare and Medicaid Services (https://data.cms.gov) was used to extract information on drugs prescribed by individual physicians and other health care providers and paid for under the Medicare Part D prescription Drug Program. Medicare Part D Claim for cocaine products including cocaine hydrochloride (HCl) and cocaine 4% was used to access medical use of cocaine. The year 2013 was selected as the earliest annum for which information was publicly available.

Data on covered outpatient drugs that are paid for by state Medicaid agencies were extracted from drug utilization databases (https://data.medicaid.gov/). Cocaine products, including cocaine hydrochloride and cocaine 4%, were considered. Both Fee-For-Service (FFS) Utilization records and Managed Care Organization (MCO) Utilization records were collected. Units reimbursed by the state during the year covered and the number of prescriptions were used to assess medical use of cocaine. FFS and MCO data were obtained using the 11-digit NDC for the volume dispensed. The number of prescriptions included any prescription for which Medicaid paid a portion or the full claim. FFS is the number of prescriptions reimbursed by the state Medicaid agency that were administered as outpatient drug claims during a given quarter or year examined. MCO is the number of prescriptions dispensed as outpatient drug claims during the quarter or year considered. These national databases were complemented with an examination of electronic medical records in the Geisinger Health System which serves over 3 million patients, primarily in central and northeastern Pennsylvania. Procedures were deemed exempt by the Institutional Review Boards of the University of New England and Geisinger.

### Data analysis

Statistical analysis was completed with Systat, v. 13.1 with *p* < 0.05 considered statistically significant. The percentage of each business activity (e.g., fraction of the total of 9,625 hospitals in 2017) that received cocaine at least once per year was determined. Heat maps were prepared with Excel with cocaine by weight (mg) per 100 persons, as determined by the annual American Community Survey. States whose use was 1.5 standard deviations above the average were considered elevated and those placing 1.96 standard deviations above were considered significantly elevated. Per capita cocaine use in 2017 was also expressed as a ratio relative to the lowest state (1.0). The number of cocaine orders was normalized to per 100K active patients in the Geisinger system (2002 = 246,379, 2018 = 556,984) with active defined as least two encounters at any time and at least one encounter in the given year. Figures were prepared with GraphPad Prism, 6.07.

## RESULTS

Total cocaine distribution per year by weight as reported by the DEA was evaluated in the United States over a 15-year span (Figure 1A). Accordingly, the highest licit cocaine usage occurred in 2002 (71.83 kg) while the lowest (26.95 kg) was recorded in 2017, indicating a 62.5% decrease. Further examination by the business activity of the recipient was completed, revealing the same pattern. In 2017, hospitals accounted for the preponderance of use (84.88%), followed by practitioners (9.88%) and pharmacies (5.23%). Cocaine distribution to mid-level practitioners or teaching institutions was very limited (< 0.02%). Similarly, the percentage of potential cocaine buyers and their distribution was also evaluated and showed declines (Figure 1B). In 2017, cocaine for medical use was stocked by less than two in five (38.4%) hospitals, one in forty practitioners (2.3%), and one in three-hundred pharmacies (0.3%; Supplemental Figure 1).

**Figure 1.**
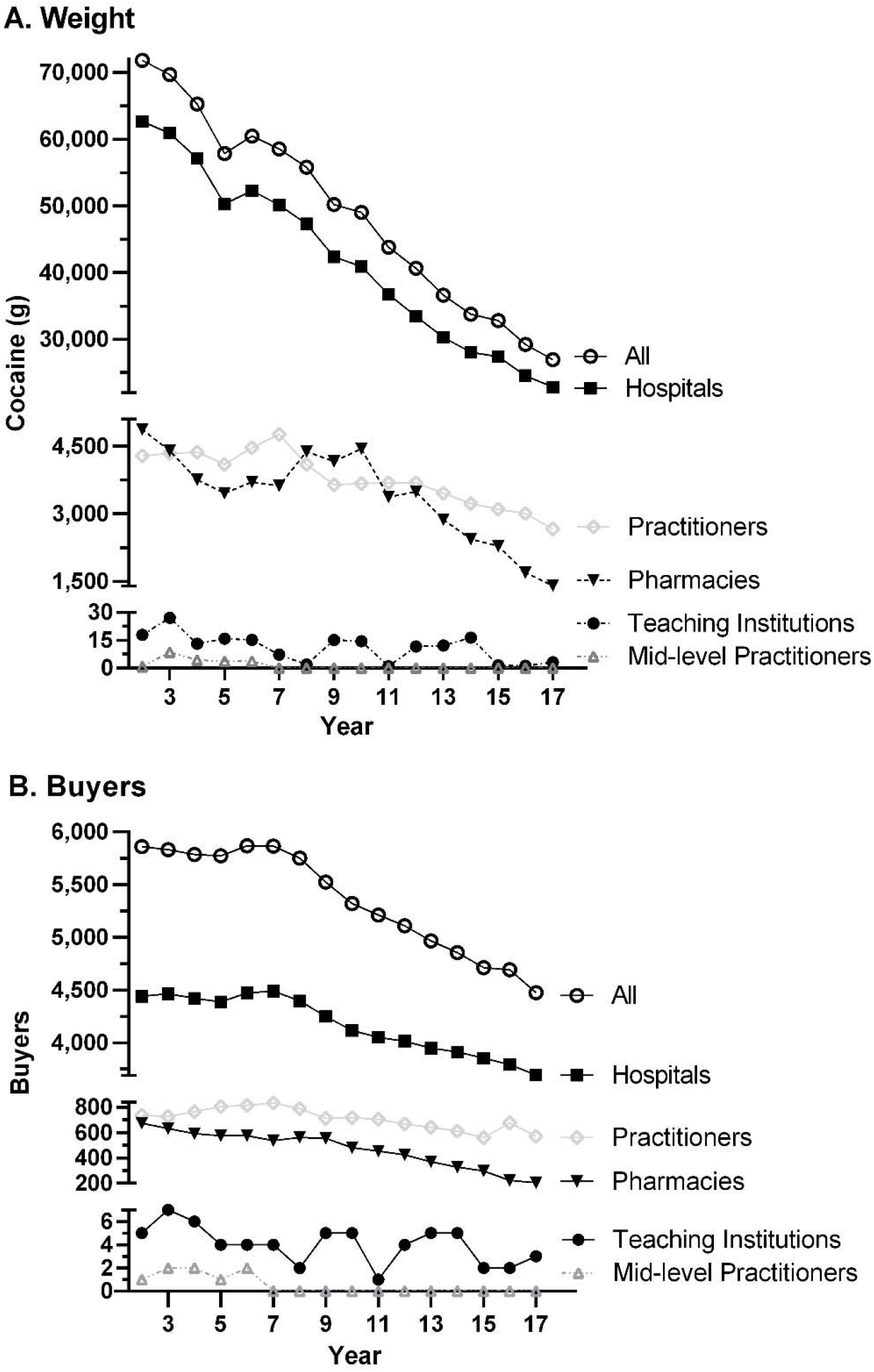
Cocaine, as reported to the US Drug Enforcement Administration, distribution by weight (top) and number (bottom) by business activity of recipient (total is the sum of hospitals, practitioners, pharmacies, teaching institutions, and mid-level practitioners) from 2002 to 2017.

When normalized for population count, pronounced regional variations in cocaine use were observed (Figure 2, Supplemental Figures 2A). There was a 7.6-fold difference between the highest use location (South Dakota = 76.1 mg / 100) relative to the lowest one (Delaware = 10.1 mg /100) states for 2002. Relative to the national average value (29.4 mg/100 persons), use of cocaine was elevated in North Dakota (> 1.5 SD above average) and significantly elevated (> 1.96 SD) in Montana, Wyoming, and South Dakota. A 10.2-fold difference between the highest (North Dakota = 24.1 mg/100 persons) and the lowest (South Carolina = 2.4 mg/100 persons/y) reporting states was observed for 2017 (Supplemental Figure 2B). Montana and North Dakota were significantly elevated as compared to the national average (9.4 mg/100 persons).

**Figure 2.**
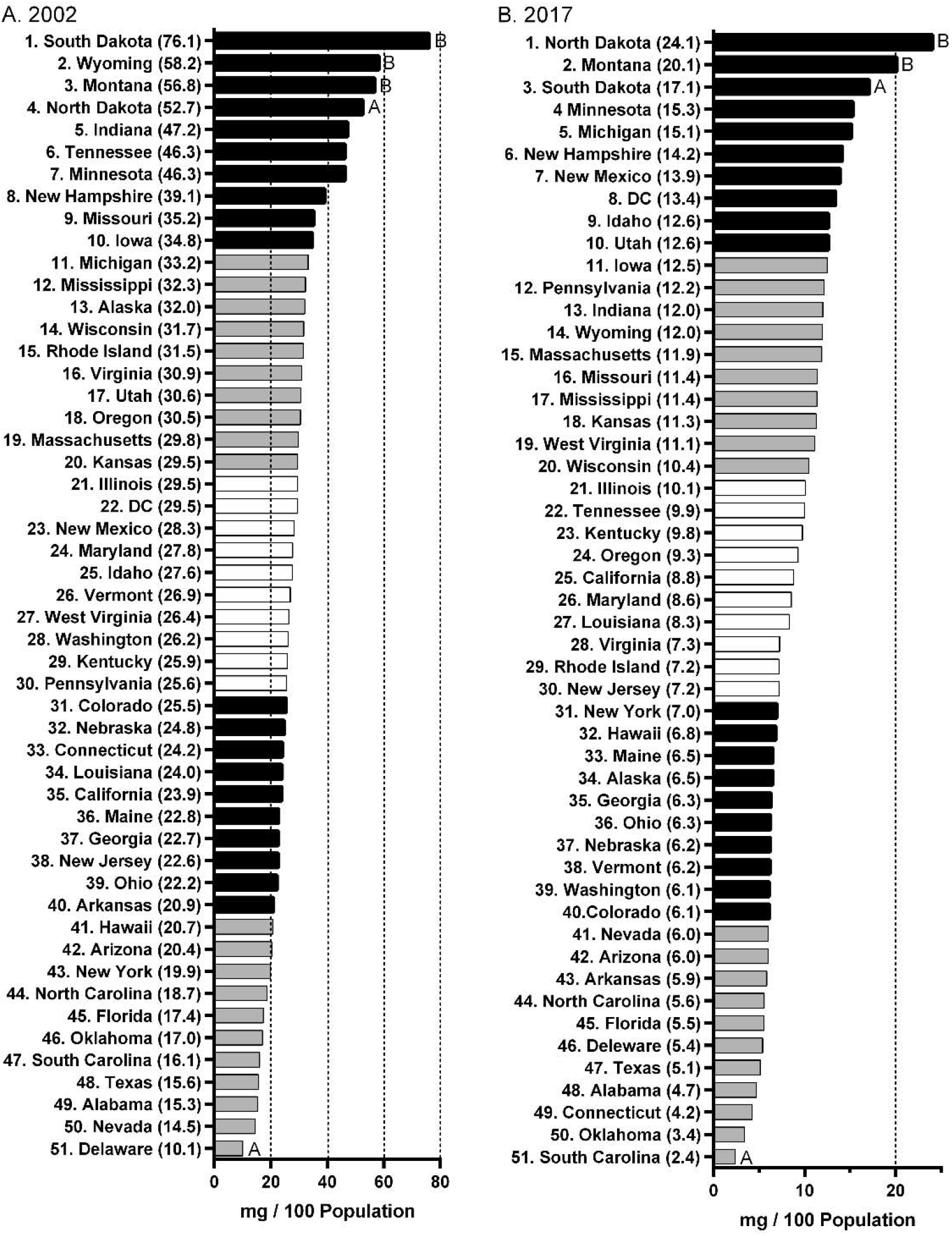
Cocaine distribution (mg) as reported to the Drug Enforcement Administration’s Automated Reports and Consolidated Ordering System, per 100 population, ranked by state in 2002 (left) and 2017 (right). District of Columbia: DC. Differs from the national average by ≥^A^ ± 1.5 or ^B^ ± 1.96 (*p* < .05) standard deviations.

Use per state decreased significantly (*t(*50) = 15.13, *p* < 0.0005). Forty-two states showed a population-normalized distribution above 20 mg/100 persons/y in 2002 versus only two in 2017. There was a high correlation in per state use from 2002 to 2017 (*r(*49) = +0.78, *p* < 0.0005). A heat map was created to depict the percent change in population-normalized cocaine distribution by state (Figure 3). South Carolina had the largest decrease (−85.3%) and Delaware (−46.4%) the smallest.

**Figure 3.**
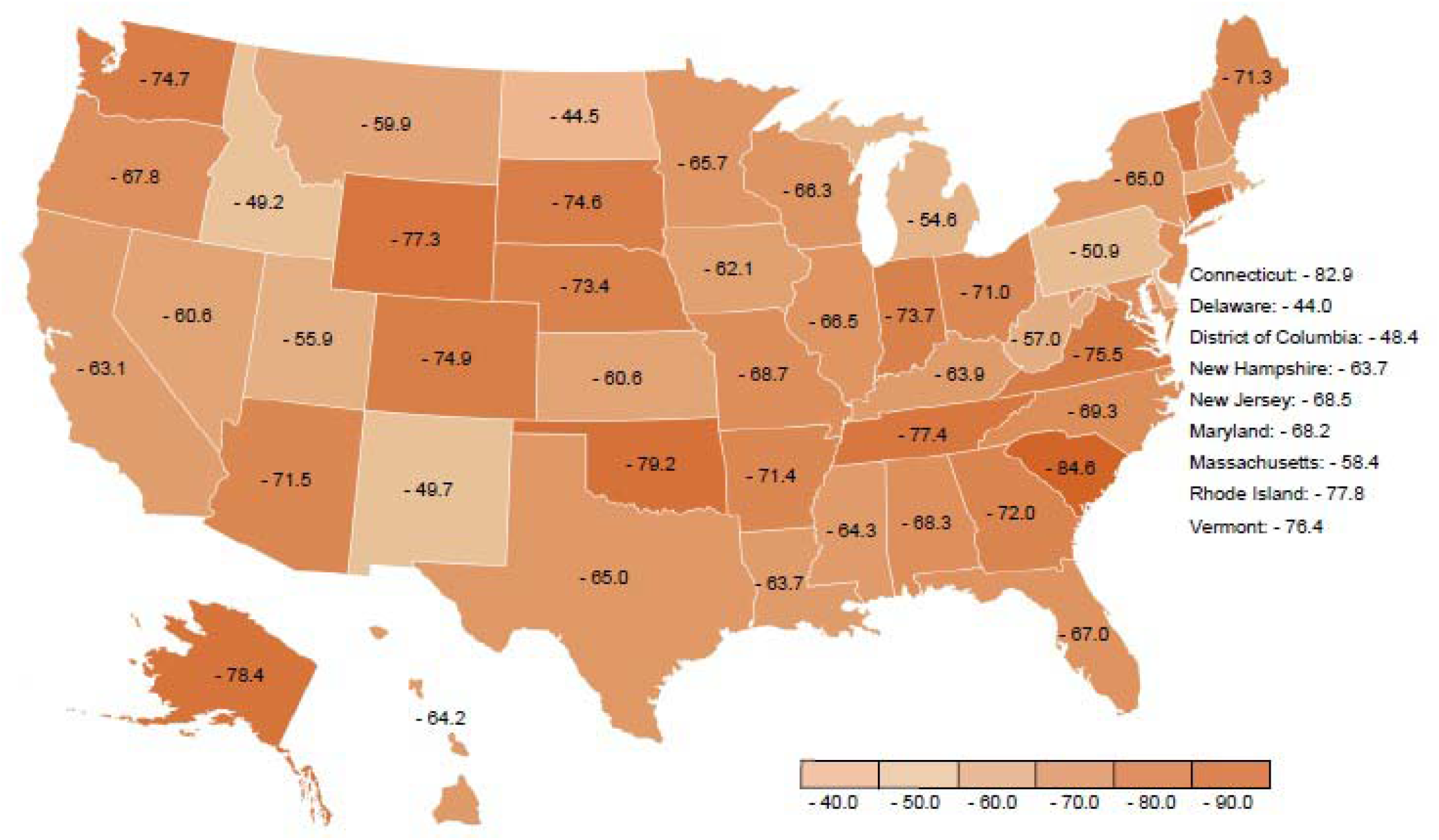
Percent reduction in cocaine distribution per 100 population by state as reported to the Drug Enforcement Administration’s Automated Reports and Consolidated Ordering System from 2002 to 2017.

Medicaid reimbursements and Medicare part D claims for outpatient cocaine procedures were evaluated from 2013 to 2017 and were extremely rare. Except for reporting year 2014, Medicare claims data had to be suppressed for all other years, because values were too low to preserve patient confidentiality.

Examination of Geisinger electronic medical records from 2002 to 2017 revealed that inpatient use accounted for the preponderance of orders (98.9%) as did the 4% solution (99.4%, N = 1,942). Additional information on the time-course showed a variable pattern (Supplemental Figure 3).

## Discussion

Cocaine distribution by weight in the US as reported by the DEA decreased substantially between 2002-2017. Similarly, the number of pharmacies carrying cocaine was determined to have declined by almost seventy percent. The primary reason for this development may be discontinued use of licit cocaine applications by physicians who previously used cocaine and an increasing number of otolaryngologists who have never elected its use in medical practice.^11^ Although some physicians still believe cocaine to be the best agent for vasoconstriction and local anesthesia, its use was discontinued due to strict regulations leading to problems with storing and dispensing this tightly controlled substance, as well as an increased availability of safer alternative medications, and prudent avoidance of its potential toxicities, which include upon application on the nasal mucosa, intraoperative hypertension and transient ventricular tachycardia.^11^ Other reported adverse cardiovascular reactions associated with intraoperative cocaine use include myocardial infarction, cardiogenic shock, cardiovascular arrest, and death. Additional reported sympathomimetic effects include mydriasis and glaucoma.^19^

Since a number of safer alternatives have become available for the medical setting and provider use, the finding of decreased medical use of cocaine was not unexpected. This decline is consistent with the phasing out of using cocaine eye drops in the diagnosis of Horner’s syndrome, a practice that was diminished starting in 1986 with reports of limitations in its diagnostic utility. One such limitation was that the cocaine eye drop test produced too many false positives in patients who did not have the disease.^20^ In 2005, cocaine was found to be a weak dilator and therefore proved ineffective for diagnosing milder forms of Horner’s syndrome (particularly partial Horner’s syndrome).^7^ Substitutions like apranoclonidine may not account for much of the observed cocaine decrease, as there is a low frequency of Horner’s syndrome in the population.^7^ Furthermore, there cannot be a complete substitution for cocaine in all cases by apranoclonidine. Infants are at risk for central nervous system depression with use of apranoclonidine, so cocaine eye drops are used instead for this population.^10^

Results showed that medical cocaine usage steady declined from 2002 to 2017. Reports for total intravenous anesthesia for head and neck procedures, propofol and opioids (i.e., remifentanil, sufentanil, fentanyl, and alfentanil) are used currently.^21^ Although propofol and opioids can induce hypotension, they facilitate intraoperative hemodynamic control and rapid recovery from anesthesia. As previously mentioned, the preferred pharmacologic agent for diagnosis of Horner syndrome in the US today is apraclonidine rather than cocaine. One of the first topical anesthetics for children, tetracaine-adrenaline-cocaine (TAC), although safe if used correctly, has been replaced by other topical anesthetics, such as lidocaine-epinephrine-tetracaine (LET), due to systemic toxicity such as hyperexcitability, seizures, stroke, cerebral hemorrhage, tachycardia, arrhythmias, malignant hypertension, and cardiac arrest.^22^ Furthermore, topical use of a solution of bupivacaine-norepinephrine has been shown to be an effective alternative to TAC for local anesthesia during the repair of lacerations in children.^23^

Evidence-based medicine generally supports the observed overall decline in cocaine distribution for medical use.^11^ There appears to be subsets of providers that continue to use cocaine for inpatient procedures (Supplemental Figure 3). However, an almost eleven-fold difference between use, normalized for population count, in the highest and lowest ranking states in 2017, may be inconsistent with evidence-based practices. For comparison, there was a threefold difference among states in use of fentanyl^12^ but twenty-fold difference for buprenorphine.^14^ Even more pronounced disparities in opioid use have been observed when considering the US Territories.^15^ Other factors like the average age of the otorhinolaryngolists or anesthesiologists in each state, and whether their training included cocaine, may be more important considerations accounting for this non-homogenous cocaine use. Practitioners in large rural areas,^24^ or in states that do not have their own medical schools, may be resistant to adopt new practices. As use in teaching institutions also declined, it may be anticipated that this will result in a medical generation that is unacquainted with the therapeutic or diagnostic utility of this agent.

The prevalence of substance use disorder is slightly higher among health care providers than in the general public.^25^ An early advocate for the medical properties of cocaine was Sigmund Freud.^26^ A recent report by a team of otorhinolaryngologists noted that although Dr. Freud was a dedicated cigar smoker, his sixteen-year survival after his first surgery indicates it was unlikely that he had oral cancer. He may have been using cocaine to self-medicate after discomfort resulting from the thirty oral procedures he underwent between 1923 and 1939. Ironically, cocaine can be responsible for oral lesions by destroying the oral lining and support tissues.^4^ Addiction to morphine and cocaine may have impacted the founder of US surgical residencies.^3^ Perhaps one upside to cocaine’s declining use is that, relative to alcohol or opioids, is that this stimulant is only sporadically reported among anesthesiologists with a substance use disorder.^25,27^

In addition to this pronounced decrease in the overall usage of cocaine, Medicare and Medicaid showed minimal prescribed usage of medical cocaine compared to total cocaine use. The total cocaine quantity prescribed by Medicare and Medicaid in 2017 was 1.6 grams or 0.0059% of the total medical cocaine usage in that year. Outpatient use of cocaine is functionally almost non-existent. Although recreational use of cocaine continues and may even be undergoing a resurgence in the US,^28^ the volumes required for confirmatory testing by analytical chemists are essentially negligible. Similarly, the amount of cocaine employed as a radioligand in Positron Emission Tomography^29^ is minute. In addition to hospital use for medical purposes, ARCOS also reports on the not insignificant volume that is distributed to researchers^30,31^ whose use is anticipated to continue.

This study had limitations. Although ARCOS is a comprehensive data source, it reports only on distribution, not necessarily on actual medical use. The amount of cocaine that is delivered to hospitals and pharmacies is reported but the quantities that expire or are subsequently misused (e.g., lost to theft) are not publicly available. The 9041L code does not allow for differentiation of cocaine subsequently administered by health care providers (e.g., 10% solution of 9041L005). The “provider” category includes not only MD and DO trained physicians but also veterinarians who may elect to use only a portion of the total cocaine accounted for, when performing diagnostic tests on small animals.^8^ The number of reimbursed prescriptions, as reported by Medicaid and Medicare, provide a clearer index of outpatient use of the drug in human patients and was negligible.

Taken together, and despite the noted limitations, this data clearly indicate that cocaine, on a national scale, is becoming extremely uncommon for use in outpatient procedures and also demonstrates its decline for inpatient uses ^11^. Continuation of this trend could result in this stimulant becoming an obscure pharmacological relic of interest only to analytical chemists, addictionologists, veterinarians, and medical historians.

## Data Availability

The Drug Enforcement Administration's data is publically available.

https://www.deadiversion.usdoj.gov/arcos/retail_drug_summary/index.html

## Acknowledgements

Jove Graham, PhD and the Center for Pharmacy Innovation and Outcomes contributed to this project.

---

**Supplemental Figure 1.**
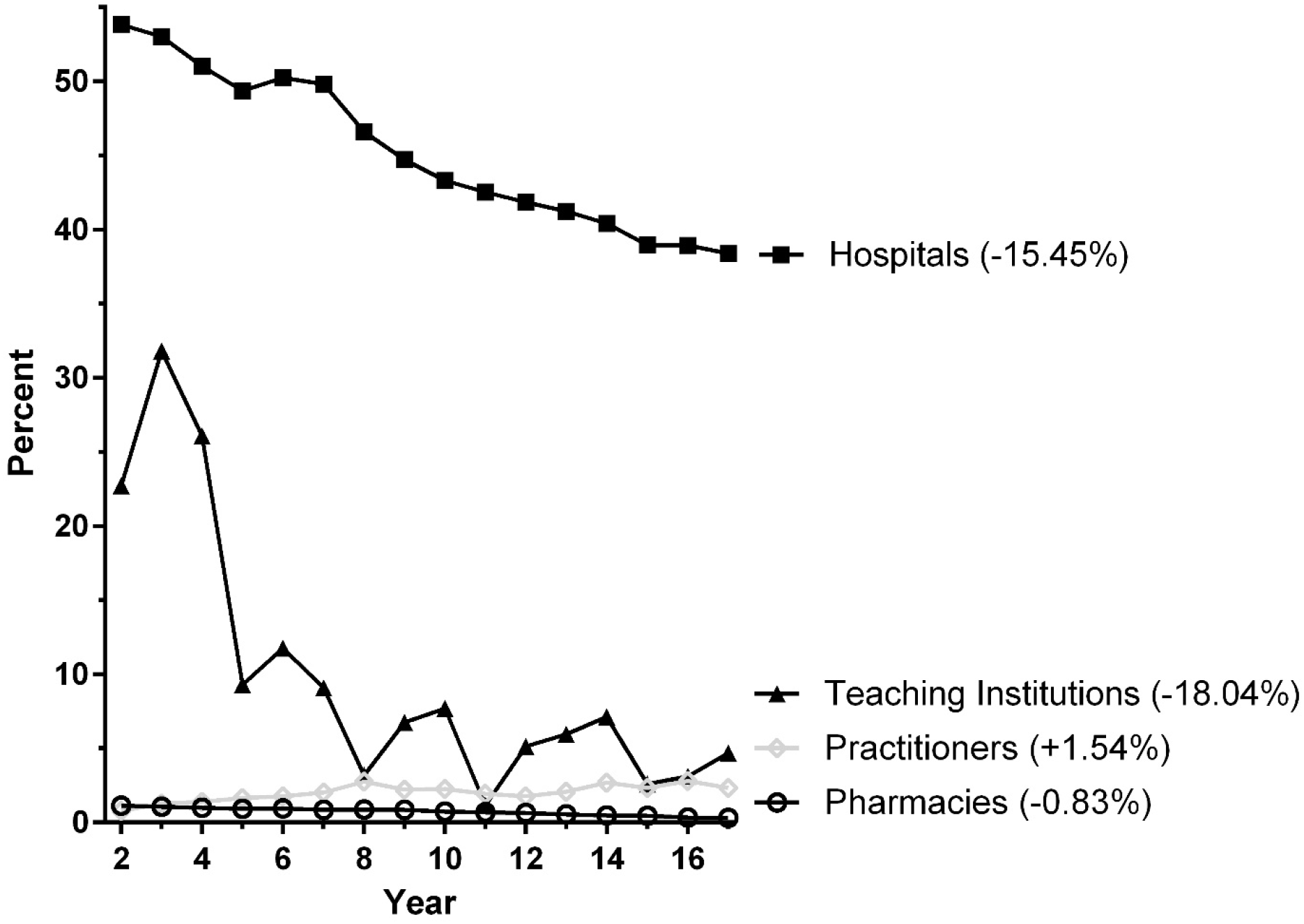
Percent of all hospitals, pharmacies, practitioners, and veterinary teaching institutions that recevied cocaine from 2002 until 2017 as reported to the Drug Enforcement Administration’s Automated Reports and Consolidated Ordering System. The demonimator was the number that received any reported controlled substance in that year. Percent change from 2002 to 2017 is shown in parentheses.

**Supplemental Figure 2.**
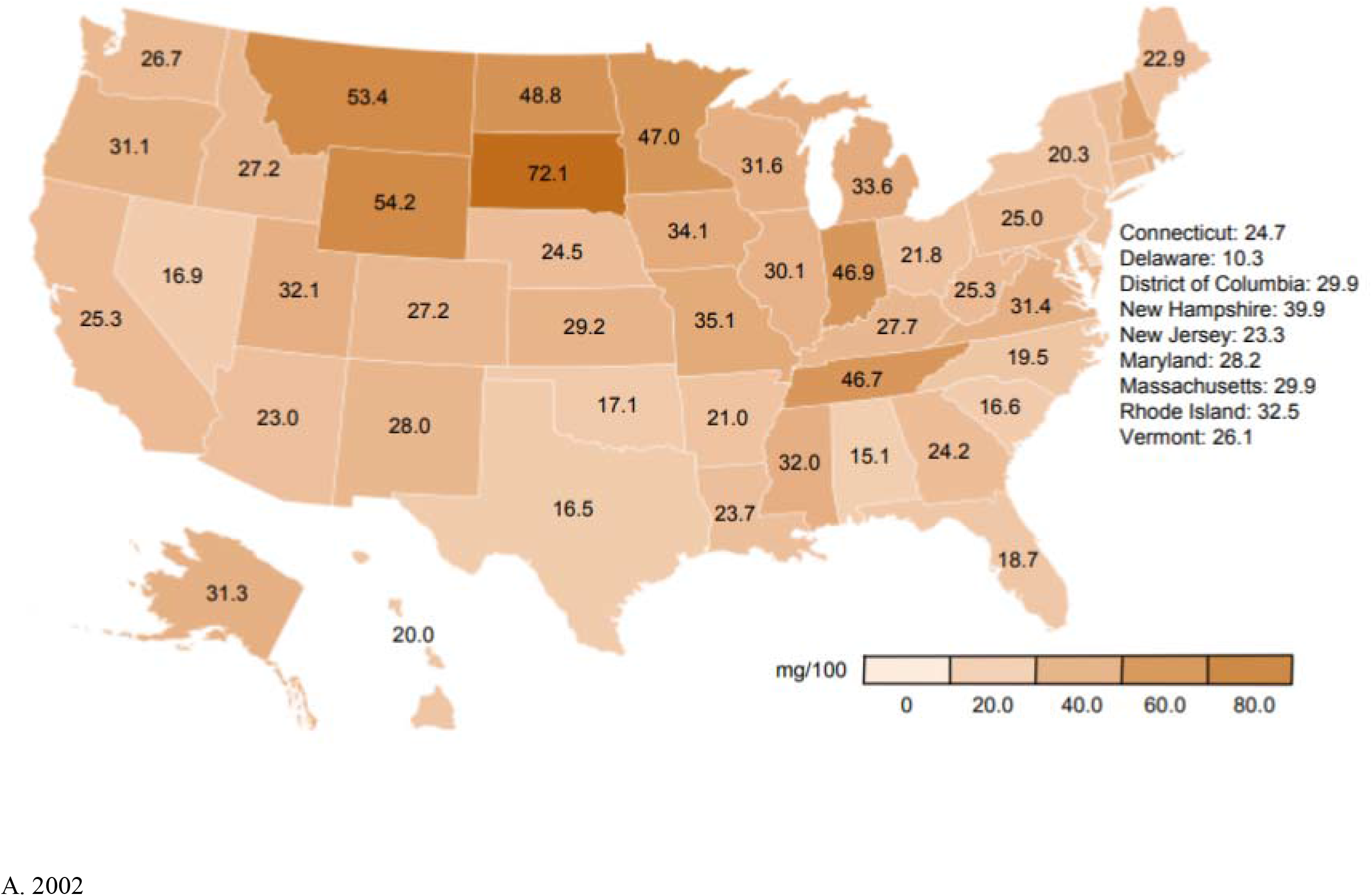
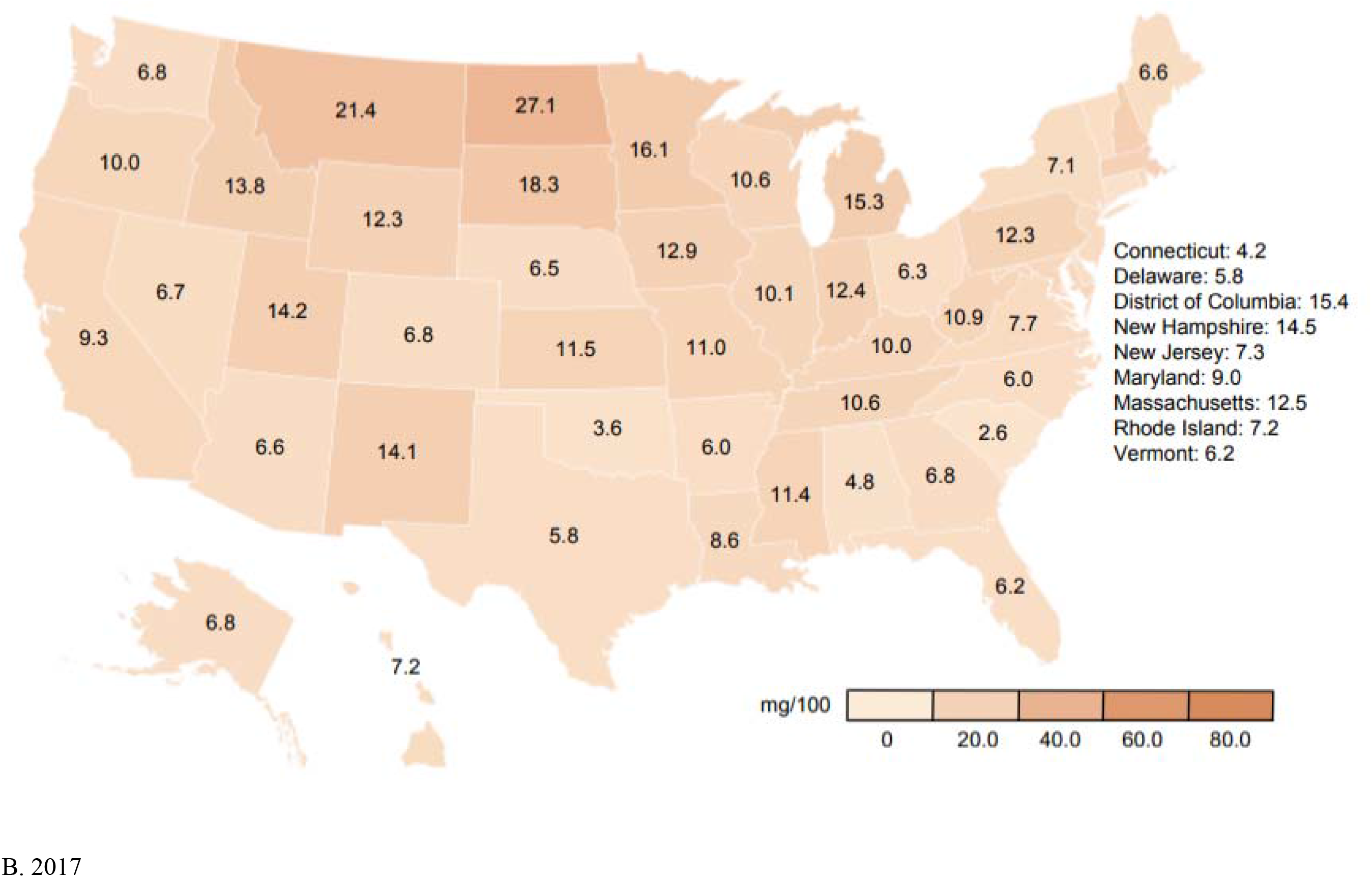
Cocaine, as reported to the Drug Enforcement Administration’s Automated Reports and Consolidated Ordering System, corrected for population (mg per 100 persons), in 2002 (A) and 2017 (B).

**Supplemental Figure 3.**
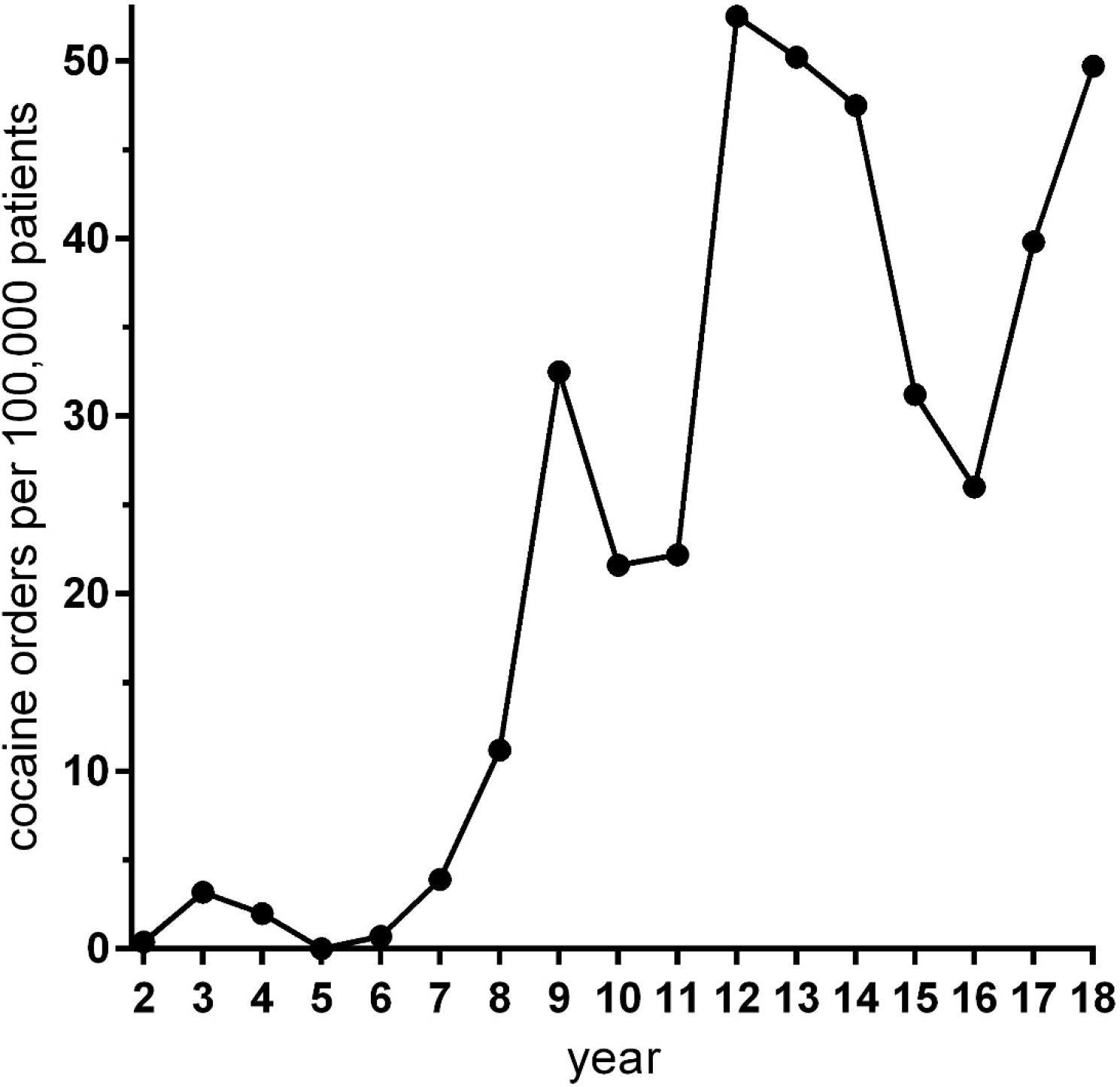
Cocaine orders per 100,000 active patients in the Geisinger Health System from 2002 to 2018.

**Supplemental Table 1.** Examples of the 125 products and formulations with the 9041L (levorotatory) code as reported by the Drug Enforcement Administration at: https://www.deadiversion.usdoj.gov/arcos/ndc/ndcfile.txt

| <u>National Drug Code</u> | <u>Ingredient</u> | <u>Trade/Product Name</u> |
| --- | --- | --- |
| 00002003S** | COCAINE HCL PDR | COCAINE HYDROCHLORIDE |
| 000020T86** | BISMUTH SERUM & COCAINE | COCAINE |
| 00002208602 | BISMUTH SERUM & COCAINE | COCAINE |
| 000020T88** | COCAINE HCL | COCAINE HYDROCHLORIDE |
| 000020Y14** | COCAINE HCL | COCAINE HYDROCHLORIDE |
| 000020Y1402 | COCAINE HCL | COCAINE HYDROCHLORIDE |
| 000020Y1425 | COCAINE HCL | COCAINE HYDROCHLORIDE |
| 000020Y39** | ADRENALIN & COCAINE #4 | COCAINE HYDROCHLORIDE |
| 000020Y84** | COCAINE HCL | COCAINE HYDROCHLORIDE |
| 000020Y8402 | COCAINE HCL | COCAINE HYDROCHLORIDE |
| 000022088** | COCAINE HCL 65MG | COCAINE HYDROCHLORIDE |
| 00002208802 | COCAINE HCL 65MG | COCAINE HYDROCHLORIDE |
| 000022514** | COCAINE HCL | COCAINE HYDROCHLORIDE |
| 00002251402 | COCAINE HCL | COCAINE HYDROCHLORIDE |
| 00002251425 | COCAINE HCL | COCAINE HYDROCHLORIDE |
| 000022539** | ADRENALIN & COCAINE | COCAINE HYDROCHLORIDE |
| 000022584** | COCAINE HCL | COCAINE HYDROCHLORIDE |
| 000022593** | ADRENALIN & COCAINE 5.4MG | COCAINE |
| 000022597** | COCAINE HCL | COCAINE HYDROCHLORIDE |
| 00002259702 | COCAINE HCL | COCAINE HYDROCHLORIDE |
| 00002259720 | COCAINE HCL | COCAINE HYDROCHLORIDE |
| 00002259302 | ADRENALIN & COCAINE 5.4MG | COCAINE |
| 000022622** | COCAINE | COCAINE |
| 000022632** | COCAINE HCL | COCAINE HYDROCHLORIDE |
| 000045221** | COCAINE | COCAINE |
| 0000A601501 | COCAINE HCL CRYST. | COCAINE HYDROCHLORIDE |
| 0000A601508 | COCAINE HCL CRYST. | COCAINE HYDROCHLORIDE |
| 0000A601601 | COCAINE FLAKES | COCAINE |
| 0000A601604 | COCAINE FLAKES | COCAINE |
| 0000E000301 | COCAINE NF ALKALOID POWD. | COCAINE |
| 0000E000308 | COCAINE NF ALKALOID POWD. | COCAINE |
| 0000Z000101 | COCAINE HCL | COCAINE HYDROCHLORIDE |
| 000191514** | COCAINE ALK.NF POWD. | COCAINE |
| 00019151430 | COCAINE ALK.NF POWD. | COCAINE |
| 00019151434 | COCAINE ALK.NF POWD. | COCAINE |
| 000191516** | COCAINE HCL USP LARGE CRY. | COCAINE HYDROCHLORIDE |
| 00019151634 | COCAINE HCL USP LARGE CRY. | COCAINE HYDROCHLORIDE |
| 00019151637 | COCAINE HCL USP LARGE CRY. | COCAINE HYDROCHLORIDE |
| 00019151640 | COCAINE HCL USP LARGE CRY. | COCAINE HYDROCHLORIDE |
| 00019151643 | COCAINE HCL USP LARGE CRY. | COCAINE HYDROCHLORIDE |
| 00019151660 | COCAINE HCL USP LARGE CRY. | COCAINE HYDROCHLORIDE |
| 00019151662 | COCAINE HCL USP LARGE CRY. | COCAINE HYDROCHLORIDE |
| 000191520** | COCAINE HCL USP FL | COCAINE HYDROCHLORIDE |
| 00019152030 | COCAINE HCL USP FL | COCAINE HYDROCHLORIDE |
| 00019152031 | COCAINE HCL USP FL | COCAINE HYDROCHLORIDE |
| 00019152034 | COCAINE HCL USP FL | COCAINE HYDROCHLORIDE |
| 00019152037 | COCAINE HCL USP FL | COCAINE HYDROCHLORIDE |
| 00019152041 | COCAINE HCL USP FL | COCAINE HYDROCHLORIDE |
| 00019152043 | COCAINE HCL USP FL | COCAINE HYDROCHLORIDE |
| 00019152053 | COCAINE HCL USP FL | COCAINE HYDROCHLORIDE |
| 00019152055 | COCAINE HCL USP FL | COCAINE HYDROCHLORIDE |
| 00019152060 | COCAINE HCL USP FL | COCAINE HYDROCHLORIDE |
| 00019152062 | COCAINE HCL USP FL | COCAINE HYDROCHLORIDE |
| 000191524** | COCAINE HCL | COCAINE HYDROCHLORIDE |
| 000192814** | COCAINE ALKALOID | COCAINE |
| 000194542** | PERUVIAN CRUDE COCAINE W/ALK 1GM | COCAINE |
| 000543082** | COCAINE HCL 4PCT | COCAINE HYDROCHLORIDE |
| 000543083** | COCAINE HCL 10PCT | COCAINE HYDROCHLORIDE |
| 000543091** | COCAINE HCL 4% 40MG TOP.SOL. | COCAINE HYDROCHLORIDE |
| 000543092** | COCAINE HCL 10% 100MG TOP.SOL. | COCAINE HYDROCHLORIDE |
| 00054315540 | COCAINE HCL TOP. SOL 10% | COCAINE HYDROCHLORIDE |
| 000743630** | COCAINE METABOLITE STOCK TRACER | COCAINE |
| 000791923** | COCAINE HCL-D3 | COCAINE-D3.HCL |

